# Quantifying superspreading in bacterial STI outbreaks using phylodynamics

**DOI:** 10.64898/2026.08.14.26360404

**Authors:** Jordi Sevilla-Fortuny, Julia Kende, Michael Meehan, Sebastian Duchene

## Abstract

Bacterial sexually transmitted infections (STIs) pose a major global public health challenge, with *Neisseria gonorrhoeae* being of particular concern due to its persistently high prevalence and increasing antimicrobial resistance. The emergence of multidrug-resistant strains has narrowed treatment options, highlighting the importance of prevention. In this context, knowing whether there is superspreading (transmission heterogeneity) within a population becomes crucial for accurate public health measures. However, classic methods to quantify superspreading rely on dense contact tracing, and this is not always feasible. As an alternative, we can use Bayesian phylodynamic modelling to infer transmission dynamics, including superspreading. Yet modelling transmission dynamics using bacterial data remains problematic, although it is widely used for viral data. Here, we apply a multi-type birth–death model parametrised to quantify superspreading in *N. gonorrhoeae* outbreaks, estimating the fraction and relative impact of superspreaders and reproductive numbers for superspreaders and non-superspreaders. We also use a hierarchical modelling strategy with partial pooling to increase the power for detecting superspreading in each cluster. Model performance was successfully evaluated across a range of superspreading scenarios using both transmission-informed phylogenies and sequence data with phylogenetic uncertainty. Application to empirical genomic data revealed a substantial role of superspreading in *N. gonorrhoeae* transmission during the COVID-19 pandemic in Australia. These results highlight the impact of superspreading in *N. gonorrhoeae* transmission and the importance of detecting it to efficiently stop the dissemination of the disease.

## 1 Introduction

*Neisseria gonorrhoeae*, the causative agent of gonorrhoea, is a common sexually transmitted bacterial pathogen and a major public health concern worldwide. Infection can lead to serious reproductive health complications, including pelvic inflammatory disease, infertility, and neonatal morbidity, particularly when untreated [1]. The World Health Organisation estimated 82.4 million new infections globally in 2020, highlighting its substantial burden [2]. In recent years, *N. gonorrhoeae* has shown increasing rates of antimicrobial resistance (AMR), most concerningly to ceftriaxone and/or azithromycin, which are the first-line treatments. The emergence and spread of resistant strains limit available treatment options and complicate infection control [3]. These challenges highlight the importance of prevention and improved surveillance to reduce transmission and control the spread of resistant lineages.

As a sexually transmitted infection (STI), the spread of *N. gonorrhoeae* is driven by host behaviour, the use of pre- and post-exposure prophylaxis, and testing and treatment rates [1, 4]. Thus, transmission can be highly heterogeneous, with a small proportion of cases contributing much more to spread than others (i.e. superspreading) [5, 6]. Mathematical modelling approaches for STIs typically consider variation in contact networks, risk behaviours, and the biology of the strain. All of these can trigger overdispersion in transmission, potentially accelerating epidemic growth and facilitating rapid dissemination [7–10].

In the absence of detailed contact tracing, the intensity of superspreading is not directly observable and may vary between different settings, locations, and time periods, strongly influencing epidemic dynamics and the effectiveness of control measures [9]. For instance, although population-level measures such as social distancing and travel restrictions applied during the COVID-19 pandemic notably disrupted *N. gonorrhoeae* transmission [11], such interventions may become less efficient than individual-level measures (i.e. testing of risk groups and notification of sexual partners) for outbreaks with high transmission heterogeneity [4, 7]. This is especially important for *N. gonorrhoeae*, as the high prevalence of asymptomatic cases allows for ’silent’ superspreading that bypasses symptom-based isolation, making proactive testing and contact tracing essential [9, 12, 13]. Thus, accurately detecting and quantifying superspreading is essential to understand transmission and to design interventions.

Current approaches to study transmission rely on epidemiological methods such as contact tracing or case counts, which aim to reconstruct transmission chains and identify individuals with a high number of secondary infections [8, 14]. However, these methods have important limitations in the context of STIs, including incomplete contact notification, asymptomatic infections, and undetected transmission events [15, 16]. As a result, superspreading may be underestimated or remain undetected using traditional epidemiological approaches. As an alternative to case counts or contact tracing, phylodynamics leverages the imprint of epidemiological processes in the evolution of genomes to infer transmission dynamics [17–19]. These methods have been successfully used to infer superspreading in viral outbreaks [20, 21]. However, the application of phylodynamics to bacterial pathogens presents additional challenges, including lower evolutionary rates and smaller transmission clusters, which reduce genetic resolution and limit the ability to accurately infer transmission patterns [22]. For instance, regarding *N. gonorrhoeae*, with a genome size of 2.2 Mb, a clock-rate of 4.6 × 10*^−^*^6^ subs/site/yr, and an infection duration of approximately three months, we would expect only around 2.5 mutations during a single infection. Moreover, the number of detectable mutations may be even lower because recombinant regions are typically masked in traditional pipelines.

In this study, we analyse each population as the result of the interaction between two subpopulations: superspreaders and non-superspreaders. Then, we use a Bayesian phylodynamic framework to estimate reproductive numbers, fraction of superspreaders and transmission advantage of superspreaders [23]. Additionally, we analyse the different clusters jointly to overcome the limitation of low genomic resolution. By doing this, the clusters share information at the population level, and thus, there is an increase in the effective sample size and an improvement in parameter identifiability [24]. This framework allows us to study real *N. gonorrhoeae* outbreaks and the impact of the COVID-19 pandemic on superspreading. The model is available as an open-source BEAST2 [25] package.

## 2 Results

### 2.1 The superspreading model

Under a constant birth-death process with no population structure, transmission is defined by the birth rate *λ*, the death rate *µ* and the sampling rate *ψ*. However these parameters are usually rearranged to define transmission in terms of *λ*, the become-uninfectious rate *δ* = *µ* + *ψ* and the sampling proportion 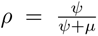. With these parameters, the basic reproductive number *R*_0_ (i.e. the average number of secondary infections in a fully susceptible population) is defined as 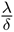. When we include population structure and use the multi-type birth-death (MTBD) model, we effectively have subpopulations, each of them with their own birth-death parameters but also with additional parameters for the interactions between demes (i.e. populations): migration rates (*m_ij_*) and birth rates or reproductive numbers between demes 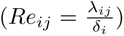. In this work, we implement superspreading in a Bayesian phylodynamic framework as a MTBD process, as described by [21, 26]. Under this implementation, superspreading is defined as a discrete trait resulting in two distinct demes: superspreaders (ss) and non-superspreaders (ns). Then, given a shared *δ*, the superspreading dynamics are defined using three parameters defined in equations 1,2 and 3: The overall reproductive number (*R_e_*, which incorporates changes in the transmission rate due to interventions or susceptible depletion), the fraction of superspreaders (*f_ss_*), and the transmission advantage of superspreaders relative to non-superspreaders (*T_a_*) respectively.

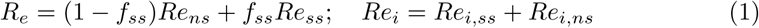

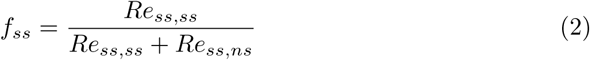

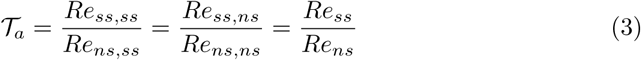

With these definitions, we can then recover for a set of *R_e_*, *f_ss_* and *T_a_* the reproductive numbers that define the MTBD as follows:

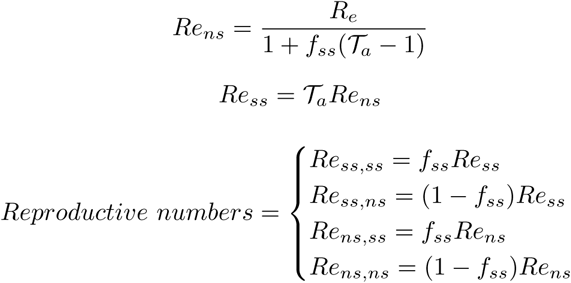

Importantly, we assume that no contact tracing data are available, and thus, we cannot assign a type to each isolate (”superspreader” or ”non-superspreader”). Instead, the type of each isolate is treated as unknown, and we integrate over their uncertainty across the two types. However, this approach leads to unidentifiability with respect to permutation of the mapping between types and rates. To avoid this problem we constrain *T_a_* to be greater than 1, implying that *Re_ss_ > Re_ns_*. Another important assumption of this parametrisation is the non-assortativity of the population. This means that the ratio at which superspreaders and non-superspreaders are born is independent of ancestry [27].

### 2.2 High levels of superspreading are accurately inferred from simulated *N. gonorrhoeae* genomes

We conducted a simulation study to evaluate the superspreading model performance for inferring the epidemiological parameters. These simulations illustrate the quality of the inferences under the particularities of *N. gonorrhoeae* data. We first simulated phylogenetic trees under different degrees of superspreading, and then used those trees to simulate sequence data. We used a *N. gonorrhoeae*-like molecular clock-rate and substitution model to account for levels of phylogenetic uncertainty similar to those observed in real data.

We analysed two types of data (trees and sequences) in BEAST2 using a novel package based on bdmm-prime [4], under two treatments (with and without deme assignment) giving rise to four different settings: (i) fixing the phylogenetic tree to the truth and providing the (correct) deme assignment to each sampled tip; (ii) fixing the phylogenetic tree to the truth and integrating over uncertainty in tip states; (iii) analysis of the sequence data (where the phylogenetic tree is inferred) and with deme assignments for each tip; and (iv) analysis of the sequence data and with no deme assignments. Setting (i) assumes that the deme of sampled individuals (superspreader or not superspreader) is known with absolute certainty and that the tree is an accurate proxy for the transmission history [28]. Setting (ii) is the situation where we have the proxy for the transmission history, but not the types of sampled individuals. Settings (iii) and (iv) are more realistic because the transmission history is unknown. In (iii) the types of individuals would be known from observed behaviour, whereas in (iv) there is only genome data available. We conducted 100 simulation replicates with four different parametrisations (see methods). For parameters estimated in each case we consider ‘accuracy’ as the number of simulation replicates for which the 95% credible interval captures the true value; ‘error’ as the difference between the true value and the posterior mean, divided by the true value (e.g. for parameter *f_ss_*, 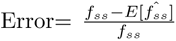 ); and ‘uncertainty’ as the width of the posterior credible interval divided by its mean. Finally, we only show in the main text the results for settings (ii) and (iv), where the tip states are unknown, as this is the most realistic scenario.

*R_e_* was accurately estimated across all simulation settings and parametrisations. When we conducted inference on fixed trees, *R_e_* was inferred with an average accuracy of 0.93 (range: 0.90 - 0.97), an average error of 0.002 (range: 0.0006 - 0.0034) and an average uncertainty of 0.12 (range: 0.10 - 0.14). Similarly, when using sequences for the inference of *R_e_*, the mean accuracy, error and uncertainty were 0.91 (range: 0.90 - 0.94), 0.01 (range: 0.0005 - 0.0180) and 0.11 (range: 0.09 - 0.15) respectively.

For *f_ss_* and *T_a_*, inferences based on sequence data, where we also infer the phylogenetic trees, were quite similar to those based on fixed trees (Figure 1), indicating that *N. gonorrhoeae*-like genomes exhibit enough genomic diversity to inform the superspreading model. However, some simulation settings resulted in more accurate inferences of superspreading. As a general rule, we expected *f_ss_* to be non-identifiable when *T_a_* approaches 1, because then the two demes are epidemiologically indistinguishable. As expected, lower values of *T_a_* led to more biased estimates of *f_ss_* (i.e. higher error, Figure 1) in our simulations. For large *T_a_*, the average error in the estimate of *f_ss_*was 0.015 (range: 0.004 - 0.039) whereas for small *T_a_* values it was 0.38 (range: 0.20 - 0.56) (Figure 1C). This effect was not evident for the inference of *T_a_*, probably due to the prior used which has most weight on low values (*P* (*T_a_ <*= 4) = 0.55). In particular, changing the treatment and specifying the states of the tips (i.e. identifying an individual as a superspreader or not) instead of integrating over their uncertainty yielded similar results overall for *T_a_*, but substantially reduced error and uncertainty in the inference of *f_ss_* (Figure S1).

**Fig. 1.**
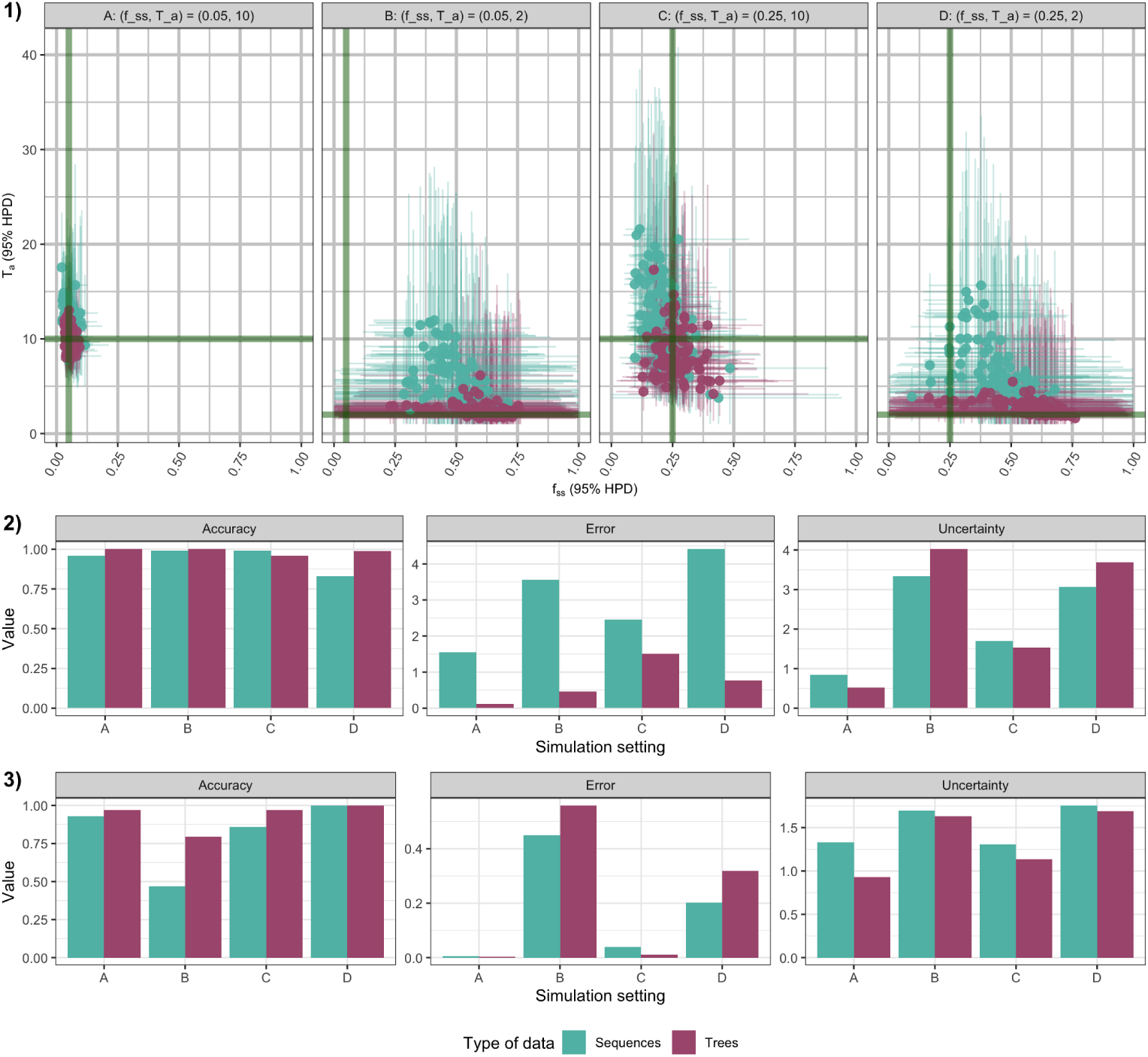
Performance of superspreading inference with different types of data and parametrisations. 1) Posterior means and 95% highest posterior density (HPD) intervals for the fraction of superspreaders (*f_ss_*, x-axis) and their transmission advantage (T*_a_*,y-axis). Panels correspond to different parametrisations (A: (T*_a_*, *f_ss_*) = (10, 0.05); B: (T*_a_*, *f_ss_*) = (2, 0.05); C: (T*_a_*, *f_ss_*) = (10, 0.25); D: (T*_a_*, *f_ss_*) = (2, 0.25)). Points represent posterior summaries, and colours indicate the data type used for inference (trees vs. sequences). Green lines denote the true simulated values. 2–3) Accuracy, error, and uncertainty of parameter estimates across parametrisations. 2) Transmission advantage. 3) Fraction of superspreaders. Colours indicate the data type used (trees vs. sequences).

We also sought to test how the model behaves when there is no superspreading, such that it is effectively overparametrised. Thus, we analysed trees simulated without superspreading under the superspreading model. Under these conditions, the model accurately recovered *R_e_*. The posterior distributions of *f_ss_* and *T_a_* were governed by the priors used on those parameters (Figure 2), a result that stresses the importance of carefully choosing priors for these parameters. Although model selection could be used in future work to formally assess the presence of superspreading, priors would still need to be selected carefully. Therefore, we used vague priors to let the data drive the inference of the intensity of superspreading.

**Fig. 2.**
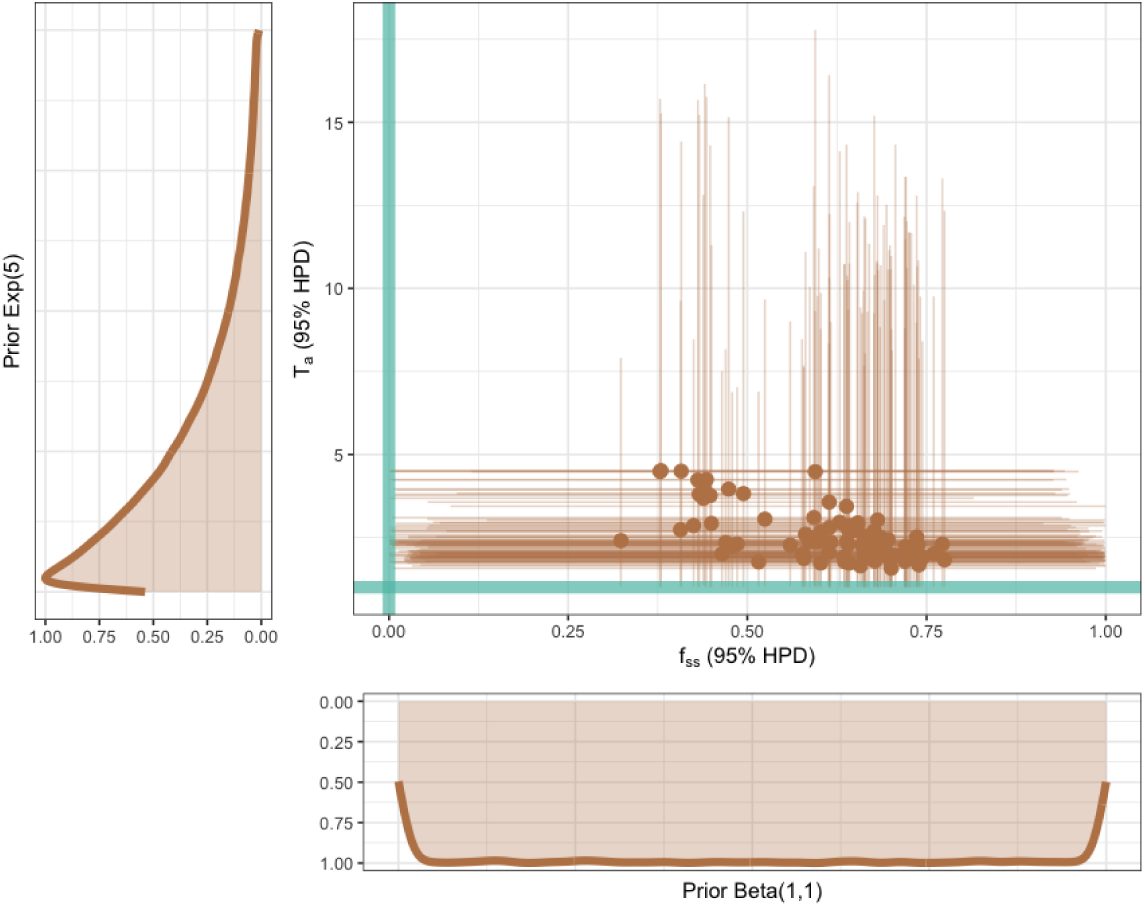
Inference under simulations without superspreading. Joint posterior estimates and 95% highest posterior density (HPD) intervals for the fraction of superspreaders (*f_ss_*, x-axis) and their relative transmissibility (T*_a_*, y-axis). Points represent posterior means and horizontal/vertical lines indicate 95% HPD intervals. Green lines denote the possible values for no superspreading. Marginal prior distributions are shown in the side panels: an exponential prior for T*_a_* (left) and a uniform Beta(1,1) prior for *f_ss_* (bottom).

### 2.3 Superspreading is widespread across *N. gonorrhoeae* transmission clusters

Once the performance of the superspreading model had been tested on simulated data, we applied it to infer transmission heterogeneity in real *N. gonorrhoeae* outbreaks sampled in the Australian state of Victoria between 2019 and 2021. Eighteen different clusters were identified with cluster sizes ranging between 33 and 528 samples (Figure S2). They correspond to putative independent introductions and circulation of this bacterium to Victoria. We analysed these clusters under a hierarchical partial pooling approach for several parameters [24]. Under this approach we analysed all the clusters within a single model. Parameters (*T_a_*, *f_ss_*, *R_e_*, *ρ* and clock-rate) are individually assigned to each cluster, and they are in turn governed by a hierarchical prior distribution (see details in Methods). The advantage of this approach over analysing clusters independently is that we allow information to be shared across clusters while preserving cluster-level variation. Furthermore, we permitted the superspreading parameters to change at the time of the first lockdown due to the COVID-19 pandemic in Australia, by introducing a skyline interval.

First of all, we assessed whether all clusters spanned a sufficient time period to observe the potential different dynamics due to the lockdown that began on 31 March 2020 in Victoria. To this end, we inspected the inferences for the origin of the process. For all clusters except for one, the origin was estimated to precede the lockdown (range: 2014-10-27 - 2020-10-10), indicating that such comparisons were feasible (Figure 3A). Importantly, the uncertainty on the inference of this parameter was high, notably for older clusters, but it is noteworthy that our inferences are consistent with such uncertainty. In particular, the only cluster for which we did not have data before the lockdown was cluster 1177, for which the origin was estimated to be the 2020-10-10 (95% HPD: 2020-02-20 - 2021-02-22) (Figure 3A).

**Fig. 3.**
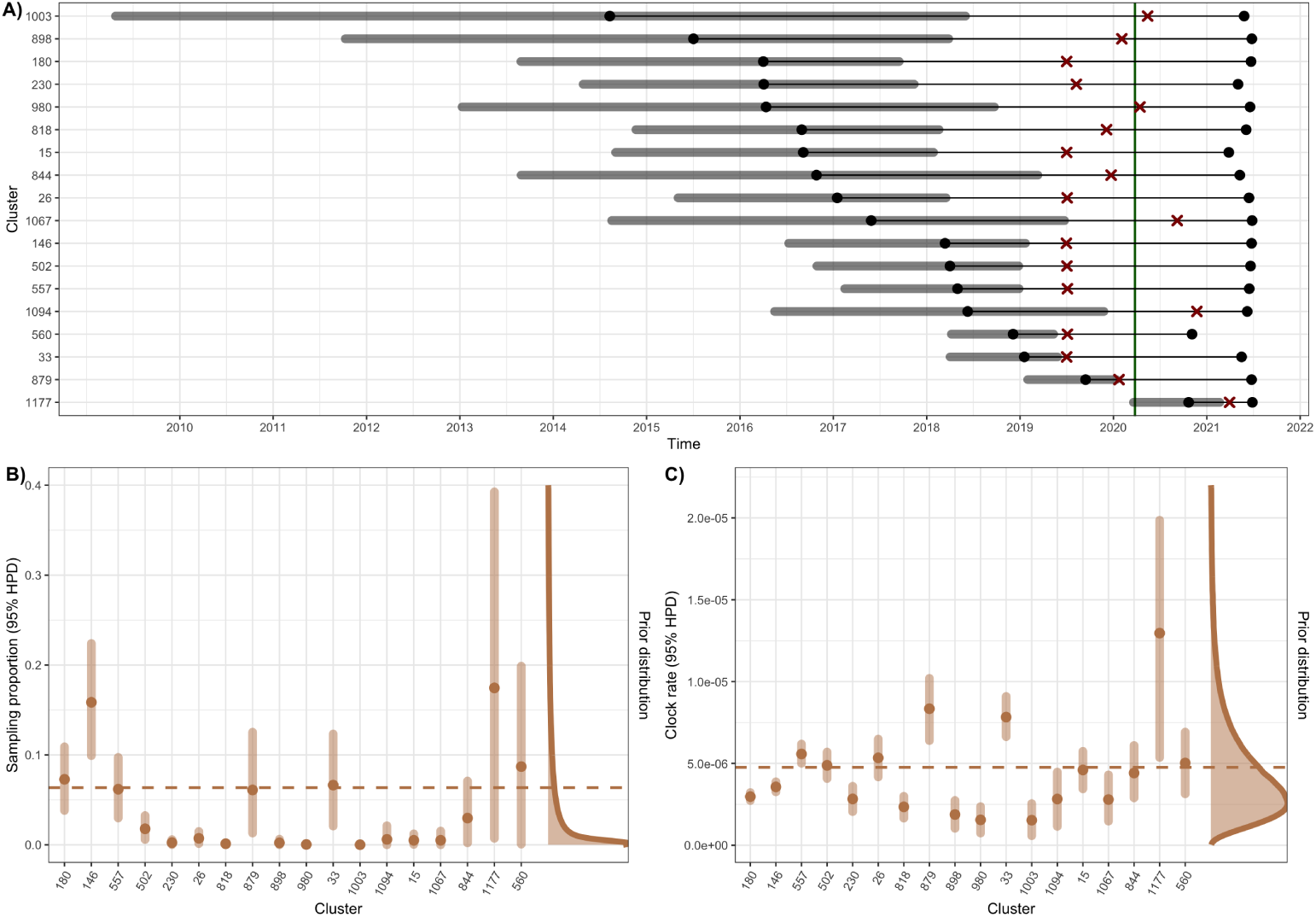
Phylodynamic inference from empirical data. A) Estimated origin times of transmission clusters. Black points at the left indicate posterior mean estimates and horizontal semi-transparent lines represent 95% highest posterior density (HPD) intervals. Red crosses denote the sampling time of the first genome in each cluster, and black points at the right correspond to the most recent sampled genome. The vertical green line marks the start of the lockdown period used in the skyline model. B–C) Cluster-specific parameter estimates under the hierarchical model. Points represent posterior medians and vertical lines denote 95% HPD intervals. B) Sampling proportion. C) Molecular clock-rate. The hierarchical prior distribution is shown on the right of each panel, with the dashed horizontal line indicating its mean.

We inspected the inferred sampling proportions and clock-rates. In both cases the partial pooling approach yielded hierarchical prior distributions consistent with previous knowledge. The posterior estimates for the *µ* and standard deviation of the log-normal prior for the clock-rate were -12.47 (95% HPD: -12.78 - -12.15) and 0.64 (95% HPD: 0.40 - 0.92) respectively. Those led to a hierarchical prior distribution with a mean of 4.68 × 10*^−^*^6^ subs/site/yr, and posterior estimates for each cluster ranging between 1.55 × 10*^−^*^6^ and 1.26 × 10*^−^*^5^ subs/site/yr (Figure 3C). Regarding sampling proportion, the posterior estimate of the shape parameter *α* of the beta distribution was 0.34 (95% HPD: 0.19 - 0.50), leading to a hierarchical prior distribution with a mean of 6.4%. Nonetheless, this distribution was heavily weighted toward low values, therefore it produced posterior estimates for each cluster ranging between 5.94 × 10*^−^*^5^ (95% HPD: 2.53 × 10*^−^*^8^ - 2.32 × 10*^−^*^4^) and 0.17 (95% HPD: 7.13 × 10*^−^*^3^ - 0.39) (Figure 3B). We wanted to test whether the low estimates of the sampling proportion in some clusters were due to the MCMC getting stuck at those low values. We reran the analysis while constraining the sampling proportion to be greater than 0.005 (0.5%), and the results were consistent with those previously described (Figure S4). Therefore, we retained the original unrestricted analysis for all downstream analyses.

With respect to transmission, we observe a decrease of *R_e_* after the implementation of the lockdown measures, as expected (Figure 4A). We calculated the posterior probability of *R_e_* being larger before the lockdown for each cluster, with an average of 0.77 (range: 0.13 - 1). Indeed, before the lockdown, all clusters were estimated to be growing (*R_e_ >* 1) with mean estimates for *R_e_*_1_ (reproductive number before the lockdown) ranging between 1.17 and 1.89. However, after the lockdown, five clusters (27.8%) switched to a non-growing scenario (*R_e_ <* 1) (Figure 4B). Notably, our estimates for *f_ss_* and *T_a_* suggest that superspreading plays a key role for the dissemination of some clusters. The estimates for *T_a_* were consistent across different clusters and time slices (Figure 4A&B), following the hierarchical prior which is a gamma distribution with mean 29.36 (95% HPD: 19.50 - 40.56). Conversely, for *f_ss_*we did observe variation between clusters and time slices (Figure 4B). In fact, we studied the presence/absence of superspreading by computing the Bayes factor for *f_ss_ >* 0.1% and considering values greater than 5 as ‘substantial’ evidence for superspreading [29]. Importantly, the hierarchical prior for *f_ss_* penalised superspreading because it was a beta distribution with beta of 5 and alpha of 0.42 (95% HPD: 0.22 - 0.63) which has more than 50% of the weight below 0.05. Using this method, we identified four different patterns of transmission: four clusters exhibited persistent superspreading regardless of the lockdown, seven showed no evidence of superspreading at any time slice, for five others there was evidence for superspreading only before the lockdown and in two clusters superspreading emerged only after the intervention (Figure 4C).

**Fig. 4.**
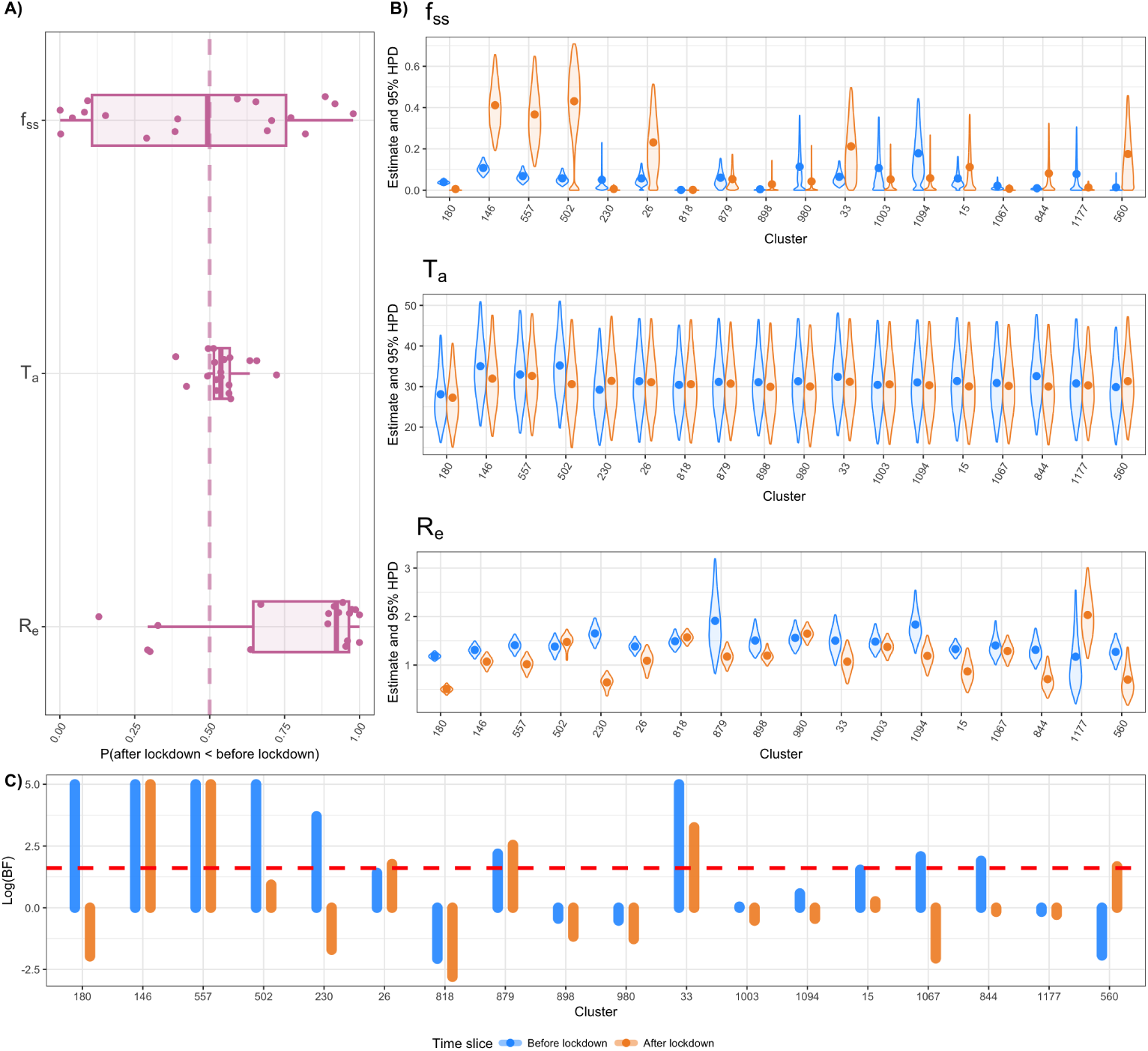
Phylodynamic inference of the superspreading parameters. A) Posterior probabilities of a decrease after the lockdown for each parameter (*f_ss_*, T*_a_* and *R_e_*). Each point is a different cluster and the pink dashed line marks the 0.5 probability (i.e. no change). B) Estimates before (blue) and after (orange) the lockdown for the superspreading parameters. Points correspond to the posterior means and the violins to the distributions for the 95% higher posterior density (HPD) intervals for each cluster (x-axis). C) Bayes factor (BF) in log scale for *f_ss_* being larger than 0.1% before (blue) and after (orange) the lockdown for each cluster (x-axis). Red line indicates a BF of 5.

### 2.4 Reduced contribution of non-superspreaders in persistent clusters after the lockdown

Once we detected the presence of superspreading in the outbreaks analysed, the next step was to depict the dynamics of those clusters in which superspreading was present. To achieve this, we studied the posterior distributions for the reproductive numbers of each deme (*Re_ns,ns_* and *Re_ss,ss_*) and the between demes reproductive numbers (*Re_ss,ns_*and *Re_ns,ss_*).

Although the overall transmission decreased on average after the lockdown, the underlying transmission patterns were highly heterogeneous. When we studied the interplay between superspreaders and non-superspreaders, we identified five different dynamics. Epidemic decline was only possible when both NS and SS were declining. Conversely, epidemic growth was possible under four scenarios: both SS and NS exhibit self-sustained growth, transmission driven exclusively by either NS or SS and transmission driven by the synergy between demes (i.e. neither NS nor SS alone could explain growth). However, the posterior distributions for the different *R_e_*showed high levels of uncertainty that coincides with uncertainty in *f_ss_*. Particularly, posterior distributions of *f_ss_* for clusters 502, 26 and 1094 were bimodal. This may be explained by the model alternating between superspreading and non-superspreading states (Figure 4B). Thus, we could not assign a unique pattern to each cluster, and instead we calculated the posterior probabilities of each scenario for each cluster.

Self-sustained growth in both demes was extremely rare in our dataset, being only plausible for the growth of cluster 879 before the lockdown and cluster 1177 after, with a posterior conditional probability of less than 0.1 in both cases (Figure 5B). Interestingly, coinciding with the lower values for the Bayes factor (Figure 4C), growth for clusters 818 and 560 before the lockdown and clusters 1067 and 818 after the lockdown was most likely due to non-superspreaders (Figure 5). Then, another common pattern was an increased importance of superspreaders for the growth after the lockdown. This was the case for clusters 146, 557, 502, 33 and 26 which are the five clusters with consistent superspreading and growth regardless of lockdown measures (Figure 4; Figure 5). Finally, the synergistic growth was most prominent before the lockdown, especially for cluster 180 which was certainly growing in this way before the lockdown according to our model (Figure 5). In fact, according to our data, the drastic decrease in growth for cluster 180 after the lockdown could be explained by the loss of the link between superspreaders and non-superspreaders. This behaviour was also observed for cluster 230.

**Fig. 5.**
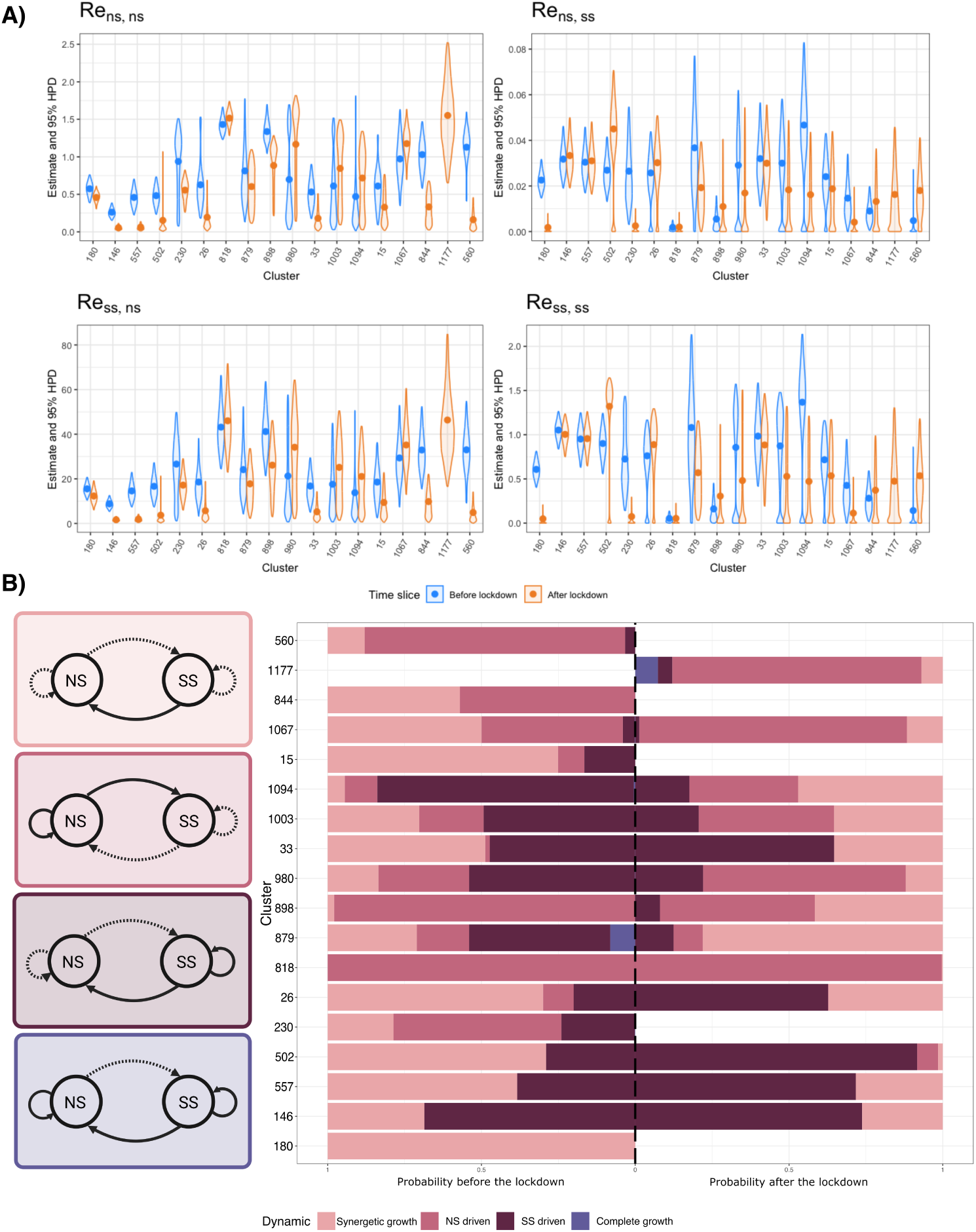
Phylodynamic inference of transmission dynamics across non-superspreading (NS) and superspreading (SS) components. A) Estimates of the effective reproduction number for non-superspreading and superspreader individuals (*Re_ns,ns_*, *Re_ss,ss_*, *Re_ss,ns_* and *Re_ns,ss_*) before (blue) and after (orange) the lockdown. Points represent posterior means, and violin plots show the 95% highest posterior density (HPD) intervals for each cluster (x-axis). B) Conditional posterior probabilities of transmission dynamics for each cluster before (left) and after (right) the lockdown. Colors indicate the relative contribution of different transmission regimes (synergistic growth, NS-driven, SS-driven, and complete growth). The vertical dashed line separates the two time periods. Schematics on the left illustrate the corresponding transmission models.

## 3 Discussion

The burden of bacterial STIs has increased dramatically in recent years especially for *N. gonorrhoeae* [2]. This increase is particularly concerning as it is coupled with high rates of AMR to first-line treatments [3]. In this context, understanding the role of superspreading is crucial for accurately characterising the pathogen’s epidemic potential and for developing more effective public health interventions. Here we present a comprehensive study on the existence, magnitude and effect of superspreading in the transmission dynamics of *N. gonorrhoeae*. Moreover, we study the effect of COVID-19-related public health measures on the superspreading dynamics. To this end, we have implemented in a Bayesian phylodynamic framework a demographic model that accounts for transmission heterogeneity [21]. As already shown by other studies, we observed a decrease in *R_e_* after the COVID-19 measures in almost all the clusters analysed. Nevertheless, the superspreading dynamics differed between clusters. In this respect, public health measures that limit spread in high-risk groups are particularly important [30].

In the Bayesian phylodynamic framework, structured models can be defined using both the coalescent and the birth-death processes [31–33]. In this study we use the structured birth-death or MTBD to define a demographic model with two demes with differential transmissibility (superspreaders and non-superspreaders). Importantly, it is usually not possible to know which samples correspond to each deme, therefore we assume unknown tip-types and integrate over the uncertainty of these assignments. That is what we tested in the simulation study conducted, where we did inference on sets of both trees and sequences simulated under different superspreading scenarios. This is important because fixing phylogenetic trees is akin to knowing the transmission history, and thus it serves as a test of correct implementation and model performance. Analyses of simulated sequence data allow us to understand the impact of phylogenetic uncertainty in the context of the information content expected in *N. gonorrhoeae* genomes. As expected, marginalising over tip states (i.e. the samples) increases the uncertainty for the inference of the epidemiological parameters, but it still yields accurate enough results especially when the levels of superspreading are substantial. Thus, this implementation permits us to infer superspreading with incomplete sampling and no contact tracing.

However, there are other limitations when we face real data, such as low cluster size that yields lower observed diversity. In fact, in the data that we analysed only three clusters have more than 100 genomes, with a median cluster size of 63 genomes. This could potentially decrease the resolution of our analysis, and thus we decided to implement a hierarchical modelling strategy with partial pooling for all the parameters for which having a common population-level distribution was plausible. This methodology has been proven to improve the inference in many different fields [34–38]. Using this strategy we also infer the hierarchical priors (i.e. the population level distributions) which can provide additional valuable knowledge. For example, in this study we obtained hierarchical priors for the sampling proportion and clock-rate whose mean values align with available knowledge [39, 40]. However, the hyperprior for the sampling proportion is strongly skewed toward very low values, indicating that, according to our model, the sampling across the analysed clusters was uneven. This suggests a few clusters were densely sampled, while most had low sampling proportions.

We found evidence for different levels of superspreading in most of the clusters analysed. These results align with previous articles describing how superspreading is prevalent in STIs [8]. However, our findings suggest that dynamics are slightly more drastic than the classic 20-80 rule (20% of cases produce 80% of infections [7]). The inference for the transmission advantage of superspreaders was consistent across different clusters with values around 30, indicating that on average superspreaders spread 30 times more than non-superspreaders regardless of the cluster. We hypothesise that the fact that we estimate a relatively constant value for *T_a_* across clusters is due to the way the model treats superspreading as a binary trait. In this case, there is likely a degree of non-identifiability between *T_a_* and *f_ss_*, as different combinations of these parameters can explain the same distribution of secondary infections. In our results, *T_a_* seems to be stable at 30 and the differences came from the fraction of superspreaders, for which the inferences were highly variable depending on the cluster. Yet, the uncertainty was substantial for some clusters making the absence of superspreaders plausible. For that reason we used Bayes factors to test for non-negligible fractions of superspreaders, and we found strong evidence for superspreading in a large number of clusters before and after the lockdown.

Finally, we studied the dynamics for each cluster before and after the COVID-19 measures. To this end we calculated the probability of growth under different scenarios (Figure 5). In a few clusters the transmission was exclusively due to non-superspreaders. However, the main pattern was a substantial contribution of superspreaders that in most cases became more important after the lockdown. This is reinforced by the fact that *Re_ns,ns_*and *Re_ss,ns_*decreased on average after the lockdown, suggesting non-superspreaders to be less involved in transmission after it. Interestingly, this aligns with previous studies showing that population-level public health measures such as lockdowns are less effective when superspreading is highly important for transmission [7]. Also, these results highlight the importance of sampling, as in most cases superspreaders are in a very low proportion but are essential for the growth of the infected population.

Although this study provides valuable knowledge about the effect of superspreading in *N. gonorrhoeae* outbreaks, there are still some caveats that have to be resolved in further work. First of all, the method developed in this study assumes that superspreading is a discrete trait, thus, it can be implemented as a particular case of a MTBD which has been already proven to be efficient and accurate. However, biologically it makes sense to think of superspreading as a continuous trait, meaning that there is a larger variance for the number of secondary infections rather than two separate demes. Then, uncertainty is still quite large for some of the inferences probably due to the low number of available genomes for some clusters. This can be partially improved using the partial pooling strategy though it stresses the need for better surveillance to increase sample sizes and new methods to optimally use the information encoded in bacterial genomes. Finally, we made valuable findings about the superspreading dynamics, nevertheless generalisation is not possible as our dataset only includes data from a very specific moment in space and time. Therefore, more datasets have to be analysed in order to accurately understand superspreading in *N. gonorrhoeae*.

## 4 Methods

### 4.1 Simulated dataset

Phylogenetic trees were simulated under varying levels of superspreading to evaluate the performance of our model for recovering the simulation parameters. A 2*^k^* factorial design was employed to define the model’s domain of applicability. *T_a_* was varied between a low value of 2 and a high value of 10, while *f_ss_*ranged from 0.05 (low) to 0.25 (high). The other parameters were held constant: *R_e_*was fixed at 1.5, the duration of infection at three months and the sampling proportion at 0.1. The simulations were performed using the BEAST2 package ReMaster [41]. For each combination of parameters, we generated 100 trees, each with 500 tips.

Sequence alignments were generated for each simulated tree to account for phylogenetic uncertainty. The simulations were done under the HKY+Γ4+I substitution model using the AliSim module of IQ-TREE 3 [42, 43]. Note that the trees generated by ReMaster are in units of time, and thus, they were scaled to genetic distance (subs/site) multiplying the branch lengths by a molecular clock-rate of 4.5 × 10*^−^*^6^ subs/site/year, consistent with estimates for *N. gonorrhoeae* [40]. The reference genome NC 011035.1 [44] was used as the seed for the evolutionary process.

Finally, another 100 trees with 500 tips each were simulated under a classic unstructured birth-death model to test the model in the absence of superspreading. The trees were simulated with the same methodology and birth-death parameters described above, except for those that pertain to superspreading, *T_a_* and *f_ss_*, which were not set (i.e. no superspreading).

### 4.2 Processing of the empirical *N. gonorrhoeae* dataset

The dataset studied by [39] was used for this research article. It includes *N. gonorrhoeae* isolates collected in Australia over a five-year period, including the COVID-19 pandemic. All the reads for samples collected after 2019 were downloaded using *fasterq-dump v3.2.1* [45]. Then, the reads were mapped against the reference genome NC 011035.1 [44] using *snippy v4.6.0* [46] with a minimum coverage of 10 reads and requiring a variant frequency of 0.9 or greater.

The sequences were classified into the transmission clusters identified in the original study. Only clusters including 30 or more sequences were used for further analyses.

A whole-genome alignment was generated for each cluster and analysed with *gubbins v3.4.3* [47] to remove within-cluster recombination.

### 4.3 Phylodynamic modelling

The epidemiological parametrisation of the MTBD model implemented in the BDMMPrime package of BEAST2 [25, 48] was used as a baseline to implement the superspreading parametrisation. This model was defined for two demes, superspreaders and non-superspreaders. Then, the MTBD parameters are defined as functions of the superspreading parameters using the feast package [49] as described in results. Importantly, for all the analyses we assumed that within each cluster the sampling proportions are the same between demes (i.e. superspreaders and non-superspreaders are sampled with the same probability) and also that there is no migration between demes, so that a change in state only occurs upon transmission.

For the simulated trees, the trees (topology and branch lengths) and the become-uninfectious rate were fixed to those used to generate them in ReMaster and to 4 *years^−^*^1^, respectively. Inference was performed only on *R_e_*, *f_ss_*, *T_a_*, and sampling proportion. Priors consisted of a lognormal distribution with mean 0 and standard deviation 1 for *R_e_*, a Beta(1, 1) distribution for *f_ss_*and the sampling proportion, and an exponential distribution with mean 5 for *T_a_*. Each simulated tree was analysed with and without specifying tip types except for the trees simulated under no superspreading, which were analysed only without specifying tip types.

Simulated alignments were included in the analysis to incorporate phylogenetic uncertainty. The epidemiological parameters were estimated using the same settings as for the inference on trees. Additionally, the tree was estimated using a clock-rate fixed at 4.5 × 10*^−^*^6^ (subs/site/year) and the HKY+Γ4 substitution model, with all the substitution model parameters jointly estimated. Each alignment was also analysed with and without specifying tip types.

Finally, a slightly more complex setting was used for the analysis of real outbreaks. First, a skyline interval was used to fix the sampling proportion before the age of the first sample to 0, thus, we only infer the sampling proportion for the interval between the first and the last collected sample for each cluster. Then, another skyline interval was employed to capture the effect of public health measures during the COVID-19 pandemic (i.e. lockdowns), permitting the superspreading parameters to change between March 20 and March 30, 2020, following the original study. Furthermore, the clusters were analysed jointly using a hierarchical structure to increase the power to detect this change. Each cluster had an independent origin of the process, but all clusters shared the same time slice for the superspreading parameters and the same become-uninfectious rate, the latter fixed at 4 *years^−^*^1^ to avoid nonidentifiabil-ity. For the remaining model parameters, a hierarchical partial pooling strategy was used, which models cluster-level parameters as draws from common population-level distributions (hierarchical priors) whose (hyper)parameters were estimated from the data via hyperpriors defined by the user (before marginalising over all other model parameters) [50]. This structure allows information to be shared across clusters while preserving cluster-level variation, inducing shrinkage toward a population-level mean and leveraging estimates for small data subsets (small trees). All priors and hyperpriors are described in Table 1. For our choice of hyperprior distributions we aimed to balance biological plausibility and a high degree of uncertainty. For example, the Beta prior on *f_ss_* reflects the fact that this parameter can only range between 0 and 1. We constrained its second parameter (sometimes known as *beta*) to 5, and for the first parameter (known as *α*) we chose a uniform hyperprior between 0 and 10. The resulting prior on *f_ss_* ranges from a skewed distribution with most density (i.e. the 95% quantile width) between 0.00 and 0.01 to one where the density is concentrated between 0.41 and 0.87. In all cases the analyses were run using 4 CPUs and sampling from the MCMC every 1000 steps. The analyses were run long enough so the effective sample sizes (ESS) for the posterior, the likelihood and the prior were above 200.

**Table 1.**
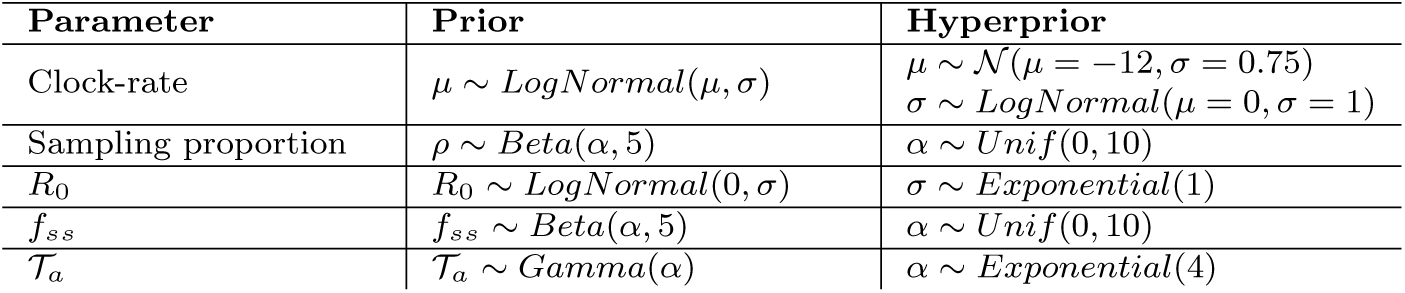
Priors and hyperpriors used for hierarchical modelling of superspreading in real *N. gonorrhoeae* outbreaks.

| Parameter | Prior | Hyperprior |
| --- | --- | --- |
| Clock-rate | $\mu \sim \text{LogNormal}(\mu, \sigma)$ | $\mu \sim \mathcal{N}(\mu = -12, \sigma = 0.75)$<br>$\sigma \sim \text{LogNormal}(\mu = 0, \sigma = 1)$ |
| Sampling proportion | $\rho \sim \text{Beta}(\alpha, 5)$ | $\alpha \sim \text{Unif}(0, 10)$ |
| $R_0$ | $R_0 \sim \text{LogNormal}(0, \sigma)$ | $\sigma \sim \text{Exponential}(1)$ |
| $f_{ss}$ | $f_{ss} \sim \text{Beta}(\alpha, 5)$ | $\alpha \sim \text{Unif}(0, 10)$ |
| $\mathcal{T}_a$ | $\mathcal{T}_a \sim \text{Gamma}(\alpha)$ | $\alpha \sim \text{Exponential}(4)$ |

### 4.4 Data analysis and visualisation

All the data analysis and figures were done in RStudio (2024.04.2+764) using R base V4.4.1 [51] and the tidyverse, beastio and ggpubr packages [52–54].

## Supplementary information

If your article has accompanying supplementary file/s please state so here.

Authors reporting data from electrophoretic gels and blots should supply the full unprocessed scans for key as part of their Supplementary information. This may be requested by the editorial team/s if it is missing.

Please refer to Journal-level guidance for any specific requirements.

## Declarations

### Code availability

The code to reproduce this study is available on GitHub (https://github.com/EDIDPasteur/superspreading manuscript). The BEAST package developed to infer superspreading with the BDMM-Prime package is also available on GitHub (https://github.com/EDIDPasteur/Beast2-SuperSpreader).

### Author contributions

M.M. and S.D. conceived and supervised the work. J.S.-F. conducted all analyses and wrote the manuscript. J.K. wrote essential computer code. All authors contributed to the writing and approved the final version of the manuscript.

## Acknowledgements

We are grateful to Tim Vaughan, who provided valuable feedback for the conception of this work.

## Ethics approval and consent to participate

Not applicable

## Materials availability

Not applicable

## Consent for publication

Not applicable

## Data availability

The accession IDs for the *N. gonorrhoeae* genomes used in this study are provided in the original publication.

