## Supplementary figures for "Quantifying superspreading in bacterial STI outbreaks using phylodynamics"

Jordi Sevilla-Fortuny[1], Julia Kende[1], Michael Meehan, Timothy Vaughan, Sebastian Duchêne[1*]

[1]Computational Biology, Institut Pasteur, Rue Du Dr Roux, 75015, Paris, France.

*Corresponding author(s). E-mail(s): ;
Contributing authors:; ; ; ;

### Supplementary File

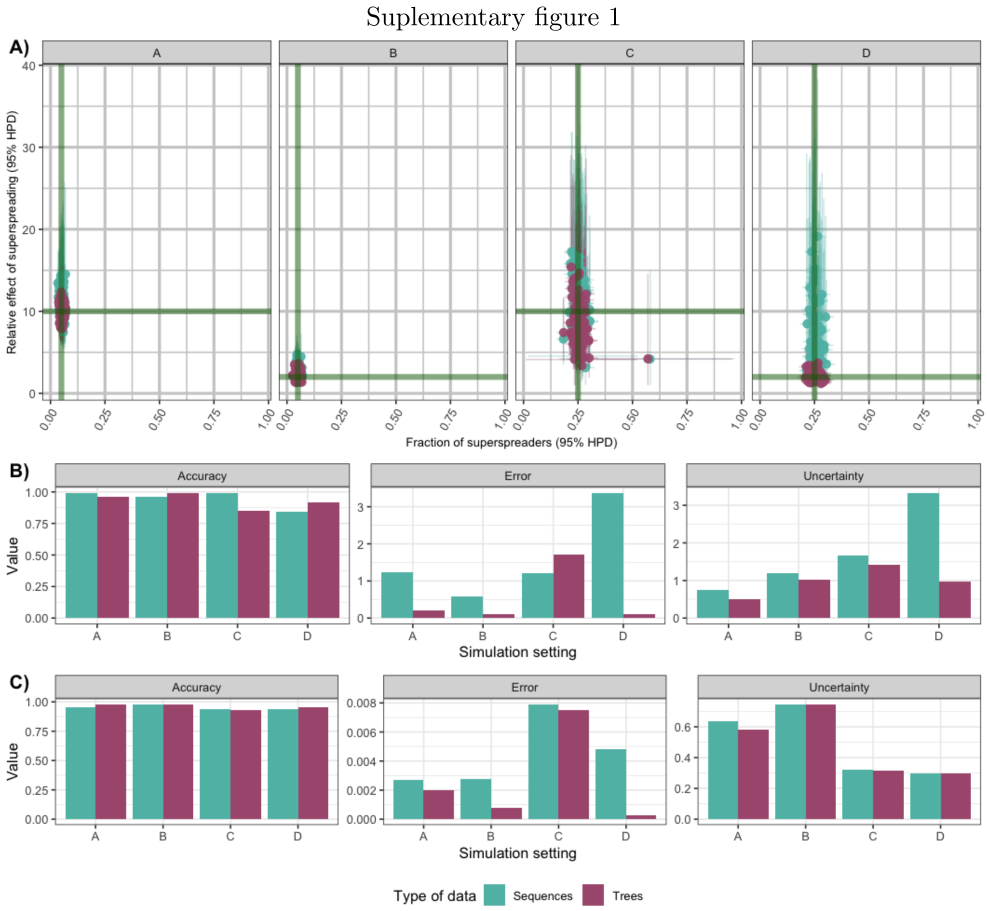


**Fig. 1** Performance of superspreading inference under varying simulation conditions with known tip-types. A) Posterior means and 95% highest posterior density (HPD) intervals for the fraction of superspreaders ($f_{ss}$, x-axis) and their transmission advantage ($\mathcal{T}_a$,y-axis). Panels correspond to different simulation settings (A–D). Points represent posterior summaries, and colours indicate the data type used for inference (trees vs. sequences). Green lines denote the true simulated values. B–C) Accuracy, error, and uncertainty of parameter estimates across simulation settings. B) Transmission advantage. C) Fraction of superspreaders. Colours indicate the data type used (trees vs. sequences).

Suplementary figure 2

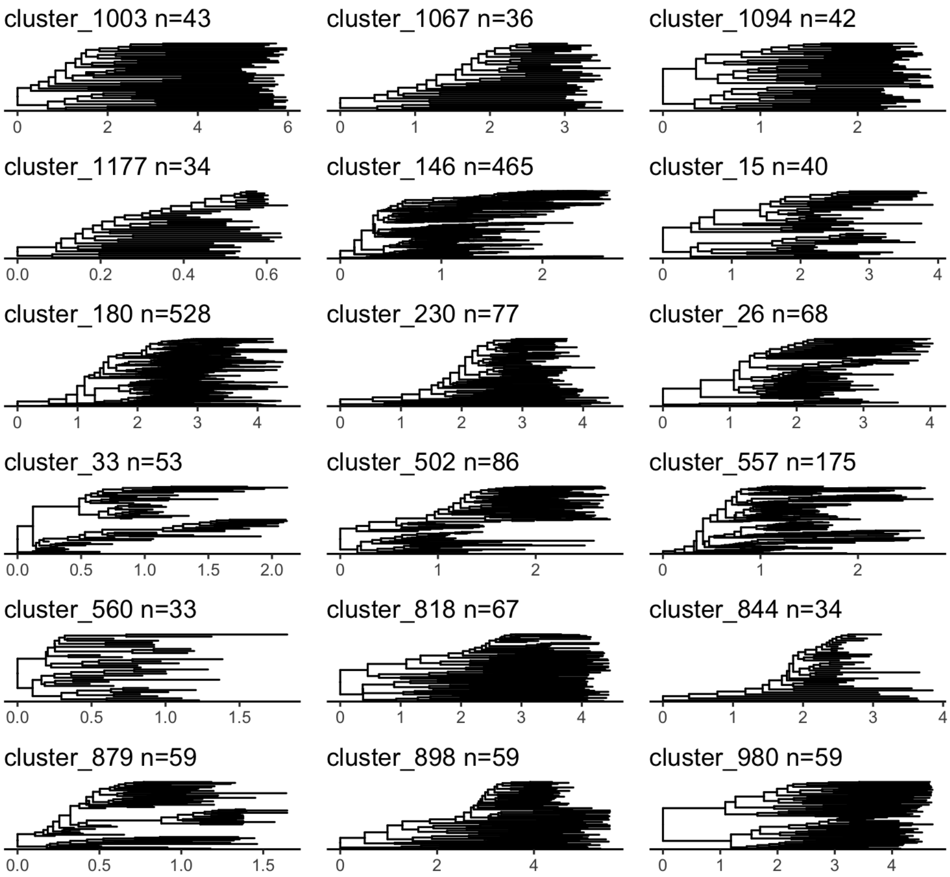


**Fig. 2** Maximum clade credibility trees for the clusters analysed. Cluster size (number of tips) is shown in the plot titles.

Suplementary figure 3

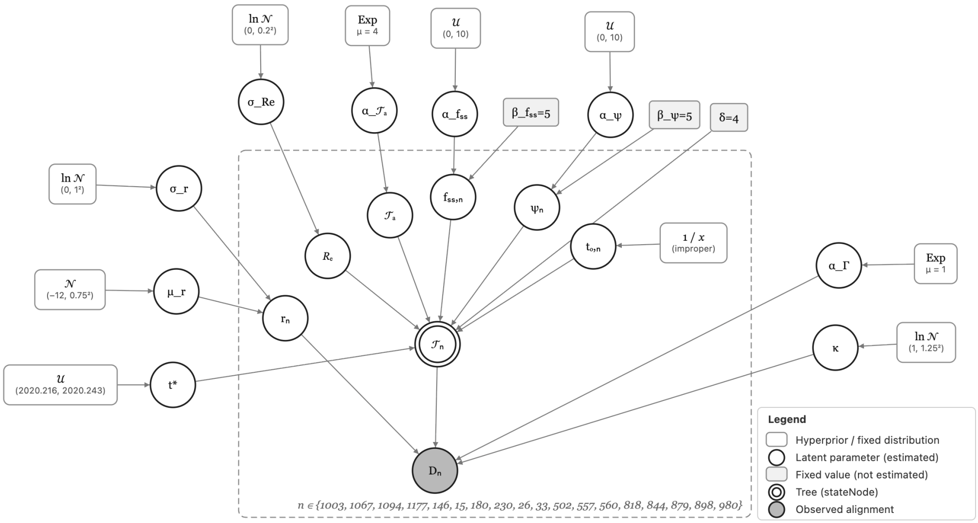


**Fig. 3** Plate diagram of the hierarchical model used to analyze the real *N. gonorrhoeae* data

Suplementary figure 4

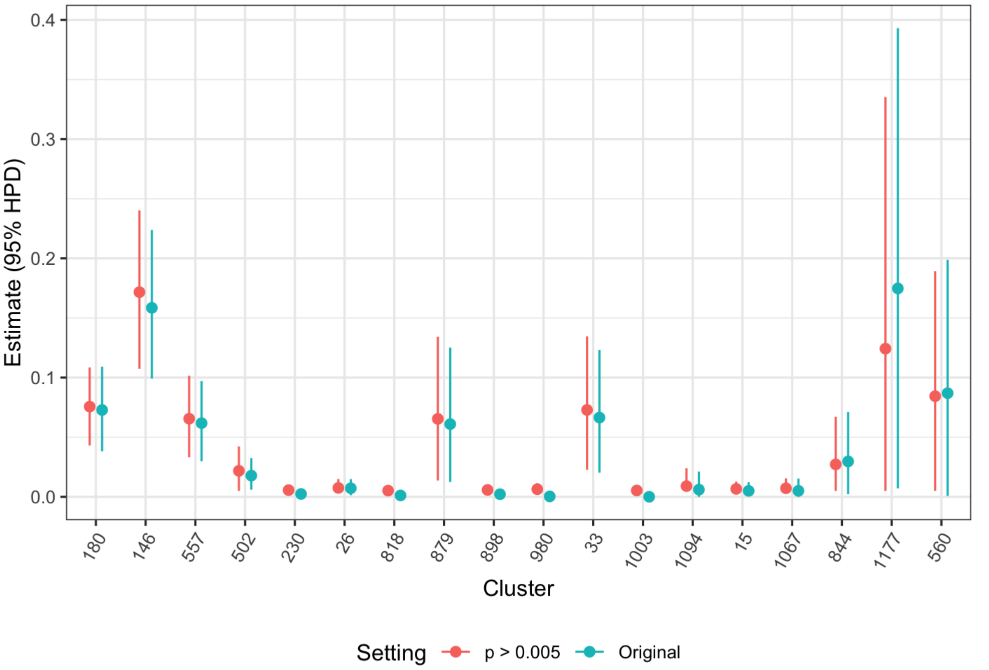


**Fig. 4** Estimates and 95% HPD intervals of the sampling proportion for each analyzed cluster. Blue points correspond to the original inference described in the manuscript, whereas red points correspond to the inference obtained when the sampling proportion was constrained to be greater than 0.005.
